# Predicting Infection-related Consultations on Intensive Care Units - Development of a Machine Learning Prediction Model

**DOI:** 10.1101/2021.03.31.21254530

**Authors:** Christian F. Luz, Dimitrios Soudis, Maurits H. Renes, Leslie R. Zwerwer, Nicoletta Giudice, Corinna Glasner, Maarten W. N. Nijsten, Bhanu Sinha

## Abstract

**Objectives:** Infection-related consultations on intensive care units (ICU) build an important cornerstone in the care for critically ill patients with (suspected) infections. The positive impact of consultations on quality of care and clinical outcome has previously been demonstrated. However, timing is essential and to date consultations are typically event-triggered and reactive. Here, we investigate a proactive approach by predicting infection-related consultations using machine learning models and routine electronic health records (EHR).

**Methods:** We used data from a mixed ICU at a large academic tertiary care hospital including 9684 admissions. EHR data comprised demographics, laboratory results, point-of-care tests, vital signs, line placements, and prescriptions. Consultations were performed by clinical microbiologists. The predicted target outcome (occurrence of a consultation) was modelled using random forest (RF), gradient boosting machines (RF), and long short-term memory neural networks (LSTM).

**Results:** Overall, 7.8 % of all admission received a consultation. Time-sensitive modelling approaches and increasing numbers of patient features (parameters) performed better than static approaches in predicting infection-related consultations at the ICU. Splitting a patient admission into eight-hour intervals and using LSTM resulted in the accurate prediction of consultations up to eight hours in advance with an area under the receiver operator curve of 0.921 and an area under precision recall curve of 0.673.

**Conclusion:** We could successfully predict of infection-related consultations on an ICU up to eight hours in advance, even without using classical triggers, such as (interim) microbiology reports. Predicting this key event can potentially streamline ICU and consultant workflows and improve care and outcome for critically ill patients with (suspected) infections.

## Introduction

Intensive care units (ICU) are hospital departments where very complex care is delivered. This complexity unfolds on multiple levels. Foremost, on the medical level, ICUs are designed for the most severely ill patients. This requires advanced medical technology for interventions, e.g. mechanical ventilation and continuous patient monitoring. The complexity in patient care is furthermore increased by the possibility that the patient’s status can quickly deteriorate. This requires prompt actions, such as the immediate administration of fluids and antimicrobials within the first hour in suspected sepsis patients [1]. On the organizational level, highly skilled ICU personnel are needed to run these units and to care for patients in continuously rotating shifts. In addition, external specialists come to the ICU for consultations. For patients with a (suspected) infection additional input on clinical microbiology, infectious diseases (ID), or antimicrobial stewardship is usually consultation-based.

The positive impact of infection-related consultations on patient care and outcome has been shown in several scenarios. A systematic review and meta-analysis of observational studies demonstrated an improvement of quality of care and reduction of mortality in patients with *Staphylococcus aureus* bacteraemia through infection-related consultations [2]. Face-to-face/bedside visits showed an improved quality of care for bacteraemic patients compared to phone-based consultations alone [3,4]. Consultations in a face-to-face or handshake approach were also found to be positively associated with quality of care in antimicrobial stewardship studies [5,6]. However, the optimal timing and trigger for infection-related consultations are difficult to set. Most often triggers for consultation are event-based (e.g., positive culture or clinical events) or at fixed time points as in the examples above.

The health state of ICU patients can change rapidly and substantially. Thus, rapid and flexible interventions are required which also includes infection-related consultations. In non-ICU patients, these requirements are much less applicable and consequently routine consultations at predefined time points are easier to implement. For ICU patients with a (suspected) infection, infection-related consultations can be initiated in different ways: i) identification of a need for a consultation by the ICU team followed by contacting the specialist (scheduled or urgent consultation); ii) identification of a need for a consultation in the microbiological laboratory, e.g. isolation of a multi-drug resistant pathogen from a patient’s specimen and notification of the respective specialist; iii) by-catch of a patient in need for consultation during an already ongoing consultation round; iv) routine monitoring of newly admitted patients by a clinical microbiologist/ID specialist. The timeliness of a consultation is a key element of high-quality consultations [7]. This can be supported and improved through process automation. Automatic notification from the microbiology laboratory to trigger ID consultations can result in a significantly decreased delay to consultation and improved quality of care [8,9]. However, while a laboratory-based approach is usually easy to implement and already part of the routine in many settings, automating the identification of patients in need of a consultation on the ICU side is more challenging.

The identification of a need for a consultation by the ICU team is based on a plethora of available information. A patient’s history, lab results, vital signs, clinical examination, clinical risk scores, and the patient’s development over time produce large amounts of data points. All of this information is taken into account by the ICU team with their expertise and experience for any clinical decision-making process. The framework of this process and its complexity is under research and the list above is far from complete [10–13]. On average intensivists make 8.9 decisions per patient per day [12]. The decision to initiate an external, infection-related consultation could, for a simplified example, be driven by several combined factors: changes in infection-related lab results such as an increase in C-reactive protein (CRP); deteriorating vital signs (e.g. increase in heart rate and decrease in blood pressure); infection-suspected results in imaging (e.g. pulmonary infiltrates in chest x-ray images); and the lack of a (timely) response to administered antimicrobial therapy. While individual clinical reasoning of ICU team members is more difficult to store in electronic health records (EHR), large numbers of data points, such as the example above, are generated at ICUs every second. This data could be used to automate, inform, and support the process of notification and triggering of infection-related consultations.

Machine learning, statistical tools to identify patterns in large amounts of data, could be ideally suited for the task to support the triggering of infection-related consultations. The use of machine learning in infectious diseases and microbiology is increasing, covers a wide range of infection-related aspects, and is often based on ICU data [14,15]. A potential utility of machine learning was established for detecting bacteraemia and sepsis or post-surgery complications [14,15]. However, the notification, initiation, or triggering of infection-related consultations has not yet been the subject of machine learning research. Therefore, this study aimed at identifying and predicting the need for an infection-related consultation in ICU patients by developing a machine learning model using data routinely collected in the EHR.

## Methods

### Study setting

This study was performed at the University Medical Center Groningen (UMCG), a 1,339-bed academic tertiary care hospital in the North of the Netherlands. Ethical approval was obtained from the institutional review board of the UMCG and informed consent was waived due to the retrospective observational nature of the study (METc 2018/081). The study included patients admitted to the 42-bed multidisciplinary adult ICU at the UMCG between March 3, 2014 and December 2, 2017, based on the use and database availability of the local EHR system during this time. Patients were included in this study if they were registered in this EHR system, did not object to their use of data in the UMCG objection registry, and were at least 18 years old at the time of ICU admission. All patient data was anonymised before analysis. The study design including the data processing is shown in Figure 1.

**Figure 1.**
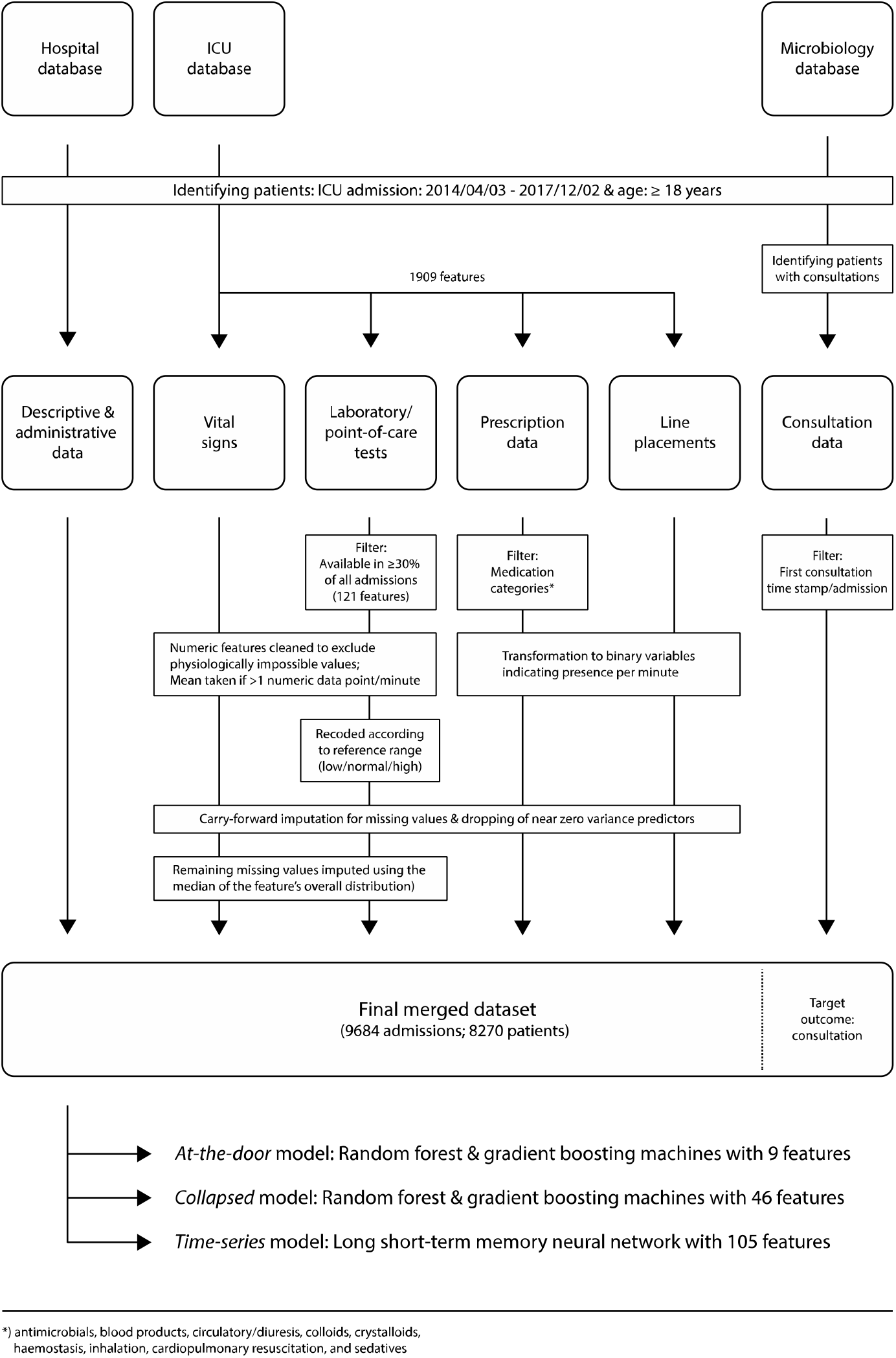
Study design and data processing for three different data sources (hospital database, ICU database, and microbiology database). Of note: (interim) microbiology reports from were not included in the modelling process. Standard data cleaning processes are not shown.

### Infection-related consultations

Consultations analysed in this study were performed by clinical microbiologists and ID specialists of the local Department of Medical Microbiology and Infection Prevention. The department is responsible for all microbiological services and hospital hygiene at the UMCG and offers a full spectrum of state-of-the-art diagnostic procedures for rapid, precise, and patient-specific diagnosis of infections. The department provides consulting assistance to the ICU in the form of a dedicated clinical microbiologist with 24/7 availability. Consultations at the ICU were triggered through different ways: 1) clinically-triggered by request from intensivists, 2) laboratory-triggered by pertinent diagnostic findings, or 3) by regular in-person participation in clinical rounds at the ICU which can also result in proactive by-catch consultations. In addition, the clinical microbiologist in charge attends the daily ICU multidisciplinary patient board. All consultations are recorded in the local database at the Department of Medical Microbiology and Infection Prevention.

### Data extraction and processing

Data was extracted from local EHR systems (Figure 1). The ICU EHR system comprised 1,909 raw features (parameters) covering all laboratory results, point-of-care test results, vital signs, line placements, and prescriptions. Demographic and administrative patient information was extracted from an administrative hospital database. Microbiological data, consultation notes, and consultation timestamps per patient were obtained from the laboratory information system (LIS) at the Department of Medical Microbiology and Infection Control. All data were cleaned to remove duplicates, extract numeric values coded in text, and standardise time stamps where appropriate.

Laboratory and point-of-care test results were included if at least one data point per feature was available in at least 30% of all patient admissions. This resulted in 121 features. Numeric features were cleaned to exclude physiologically impossible values, which typically indicated faulty tests or missing test specimens. All feature timestamps were available at the minute level. For numeric features the mean per minute was taken if more than one data point per minute was available. No double entries per minute were observed for categorical values. Categorical values indicating missing specimen or other error messages (e.g. “wrong material sent”) were transformed to missing values. Features were recoded to indicate if values fell in between feature-specific reference ranges based on available, local reference values.

Raw vital signs were cleaned for physiologically impossible values (e.g. systolic arterial blood pressure smaller than diastolic arterial blood pressure), which usually indicated faulty measurements (e.g. through kinked lines). Line placement data was transformed to a binary feature indicating the presence or absence of an intravenous or arterial line per minute and line type. Prescription data were filtered to include prescriptions of the categories: antimicrobials (identified through agent specific codes in the EHR), blood products, circulatory/diuresis, colloids, crystalloids, haemostasis, inhalation, cardiopulmonary resuscitation, and sedatives. Selective digestive decontamination (SDD) is a standard procedure in our hospital and was thus indicated by a distinct variable to avoid confusion with antimicrobial agents for other purposes [16]. All prescriptions were transformed to binary features indicating the presence of a prescription per minute, agent, and type of administration. Additional binary features were introduced indicating the presence of a prescription per prescription category and type of administration.

Missing values were filled with the last available data point. This carry-forward imputation process was used to mimic common physician’s behaviour. Remaining missing numeric values were imputed using the median of the feature’s overall distribution. Near zero variance predictors (features not showing a significant variance) were dropped from the dataset at a ratio of 95/5 for the most common to the second most common value. All data were merged per minute of ICU stay. Each patient’s stay was treated as an independent stay but a readmission feature was introduced.

Consultation data included time stamps of clinical microbiology consultations for patients admitted to the ICU. Consultation notes were available but discarded for this study. The type of consultation (i.e. via phone or in person) was not available. Data was filtered to identify the first consultation time stamp per patient and admission. This point in time formed the targeted outcome of this study. The final dataset used in the modelling process comprised 105 features (see Appendix 1).

### Cohort investigation

Descriptive analyses were stratified by consultation status (consultation vs. no consultation). Baseline patient characteristics were assessed and compared with Fisher’s exact test for categorical features and Student’s t-test for continuous features. Logistic regression was used to create an explanatory model for clinical microbiology consultations using the baseline features. Odds ratio with 95% confidence interval were used in the results presentation.

### Modelling process

Three different modelling approaches were used and evaluated in this study to predict an ICU patient receiving a clinical microbiology consultation. The first approach (*at-the-door* model) was applied using patient features available at the time of admission: gender, age, body mass index, weekend admission, mechanical ventilation at admission, sending speciality, planned admission, readmission, and admission via the operation room. Random forest (RF) and gradient boosting machines (GBM) were used to model the occurrence of a consultation.

The second modelling approach (*collapsed* model) also used RF and GBM and additional procedural features such as the presence of medication, lines, or performed diagnostics (see Appendix 1). Data was aggregated taking the mean and the standard deviation over the available time series to predict the target event (consultation). Both approaches applied a 80-20 split to the dataset to generate the training and test set. 10-fold cross validation was used to select the optimal hyperparameters of the models. The final performance was evaluated against the held-out test set.

The third approach used a long short-term memory neural network (LSTM) to model the target outcome (*time-series* model). LSTM is an artificial recurrent neural network with the advantage to include memory and feedback into the model architecture. This *time-aware* nature (and its similarity to clinical reasoning) in addition to previous reports on the beneficial use of LSTM in the field of infection management with EHR built the background for the choice of this methodology [17–19]. Using an LSTM brings the advantages that the data did not need to be collapsed to be used in the model but all available information could be used without the need for additional feature pre-processing. The LSTM model also included all available features including vital signs (see Appendix 1). For the LSTM model a 60-20-20 split to the data was applied with 60% of the data used for training, 20% for selecting the hyperparameters, and 20% for testing and reporting the model final model performance.

Model performances were evaluated and compared using the area under the receiver operator curve (AUROC) and the area under the precision recall curve (AUPRC).

All data processing and analyses were performed using tidyverse in R and numpy, pandas, caret, scikit-learn, keras, and tensorflow in Python [20,21]. The data is registered in the Groningen Data Catalogue (https://groningendatacatalogus.nl). The study followed the Transparent Reporting of a Multivariable Prediction Model for Individual Prognosis or Diagnosis (TRIPOD) statement [22].

## Results

### Patient population

In total, 8 270 unique patients and 9 684 admission were included in the study (Table 1).

**Table 1.** Patient characteristics.

|  | Consultation<br>(N=759) | No consultation<br>(N=8925) | Total<br>(N=9684) | p-value* |
| --- | --- | --- | --- | --- |
| Gender |  |  |  | 0.762 |
| Missing | 0 | 2 | 2 |  |
| Female | 290 (38.2%) | 3459 (38.8%) | 3749 (38.7%) |  |
| Male | 469 (61.8%) | 5464 (61.2%) | 5933 (61.3%) |  |
| Age at admission |  |  |  | 0.022 |
| Missing | 0 | 2 | 2 |  |
| Mean (SD) | 61.7 (14.6) | 60.3 (15.3) | 60.4 (15.3) |  |
| Range | 19 - 92 | 18 - 101 | 18 - 101 |  |
| Readmission |  |  |  | < 0.001 |
| Missing | 0 | 2 | 2 |  |
| No | 656 (86.4%) | 8249 (92.4%) | 8905 (92.0%) |  |
| Yes | 103 (13.6%) | 674 (7.6%) | 777 (8.0%) |  |
| Admission via operation room |  |  |  | < 0.001 |
| Missing | 0 | 3 | 3 |  |
| Not via OR | 508 (66.9%) | 3181 (35.7%) | 3689 (38.1%) |  |
| Via OR | 251 (33.1%) | 5741 (64.3%) | 5992 (61.9%) |  |
| Planned admission |  |  |  | < 0.001 |
| Missing | 0 | 3 | 3 |  |
| No | 635 (83.7%) | 4343 (48.7%) | 4978 (51.4%) |  |
| Yes | 124 (16.3%) | 4579 (51.3%) | 4703 (48.6%) |  |
| Length of ICU stay (days) |  |  |  | < 0.001 |
| Mean (SD) | 9.0 (10.7) | 3.6 (5.4) | 4.0 (6.156) |  |
| Median | 5 | 2 | 2 |  |
| Range | 1 - 119 | 1 - 96 | 1 - 119 |  |
| Hospital mortality (ICU) |  |  |  | < 0.001 |
| died | 348 (45.8%) | 365 (4.1%) | 713 (7.4%) |  |
| survived | 411 (54.2%) | 8560 (95.9%) | 8971 (92.6%) |  |

\*) Comparing consultation vs. no consultation; Fisher's exact test for categorical features; Student's t-test for continuous features

### Consultations

The proportion of patients with a consultation did not show a significant trend over time and centred around the mean of 7.9% (SD: 1.4) of all patients over all quarters of the study period (Figure 2). An explanatory multiple logistic regression analysis using available basic patient and administrative features was performed to identify characteristic of the consultation cohort. Several features showed a significant positive association with consultations: age, mechanical ventilation at ICU admission, and readmission (Table 2). A significant negative association with consultations was found for admission via the operation room and planned admissions.

**Table 2.** Multiple logistic regression model for clinical microbiology consultations at the ICU.

| Variable | Univariate analysis |  | Multivariate analysis* |  |
| --- | --- | --- | --- | --- |
|  | OR (95% CI) | p-value | OR (95% CI) | p-value |
| **Weekend admission** | 1.66 (1.38-1.99) | <0.001 | 0.89 (0.73-1.07) | 0.226 |
| **Gender** |  |  |  |  |
| female | reference | N/A | reference | N/A |
| male | 1.02 (0.88-1.19) | 0.757 | 1.03 (0.88-1.20) | 0.720 |
| **Age (years)** | 1.01 (1.00-1.01) | 0.022 | 1.01 (1.00-1.01) | <0.001 |
| **Mechanical ventilation at ICU admission** | 1.24 (1.07-1.45) | 0.004 | 1.67 (1.42-1.96) | <0.001 |
| **Readmission** | 1.92 (1.53-2.39) | <0.001 | 1.37 (1.09-1.72) | 0.008 |
| **Admission via operation room** | 0.27 (0.23-0.32) | <0.001 | 0.47 (0.38-0.56) | <0.001 |
| **Planned admission** | 0.19 (0.19-0.22) | <0.001 | 0.28 (0.22-0.35) | <0.001 |

OR = Odds Ratio, CI = Confidence Interval
\*) Adjusted for all variables presented in the table

**Figure 2.**
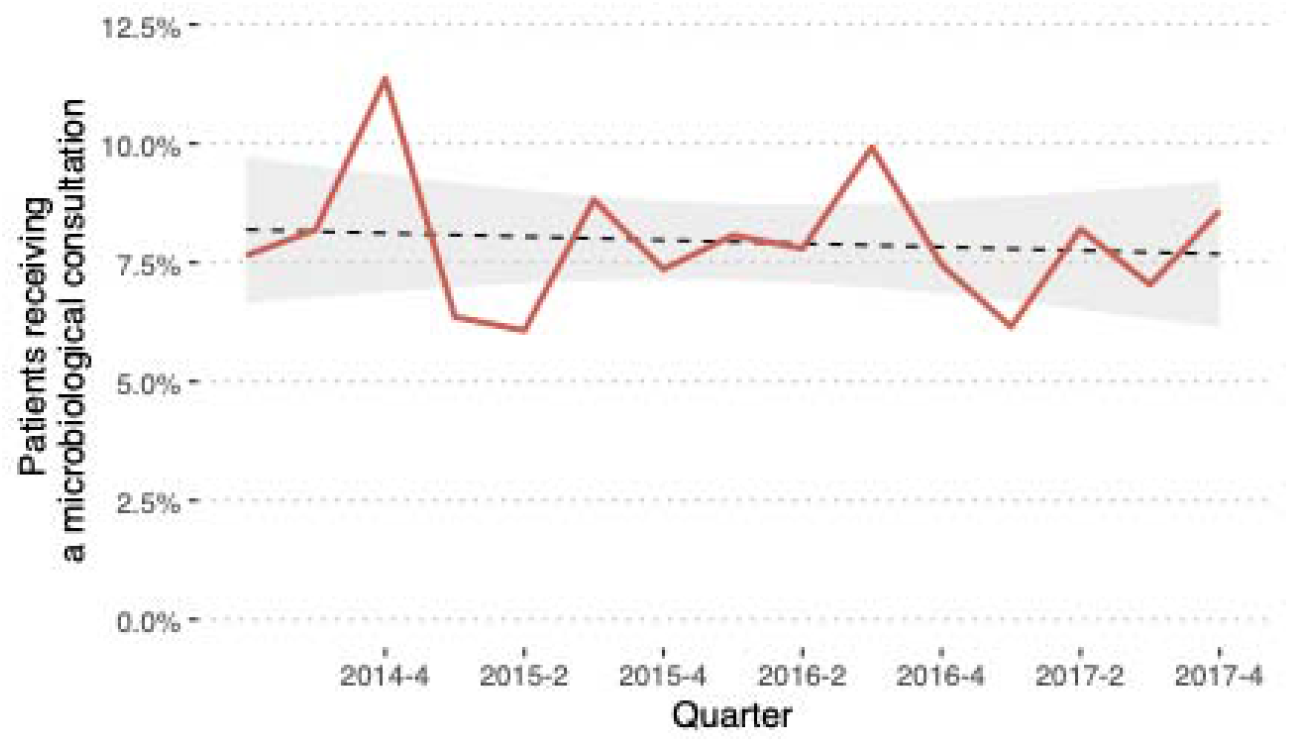
Proportion of admissions with a clinical microbiology consultation per quarter. No significant change in trend line (dashed) with 95% confidence interval (grey) using a linear regression model.

In patients receiving a clinical microbiology consultation, a significant difference in proportion of weekend admissions and weekend consultations could be observed in univariate analysis (p < 0.001) (see also Figure 3).

**Figure 3.**
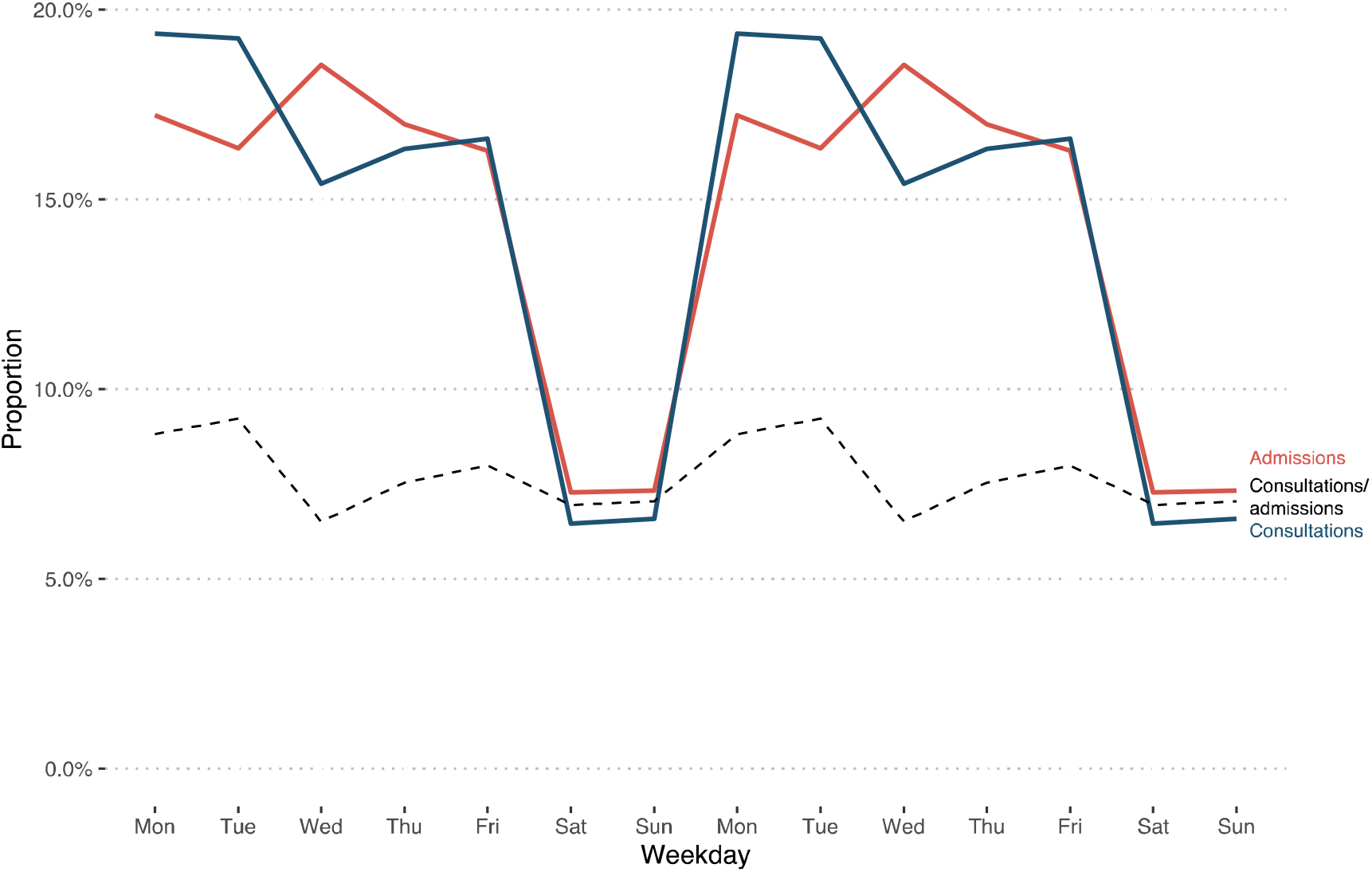
Proportion of overall admissions and consultations stratified per weekday (weekdays duplicated to display two weeks). The dashed line shows the proportion of patients receiving a consultation among all admissions, stratified per weekday.

### Predicting consultations

The developed prediction models targeted at predicting a consultation reached moderate to high diagnostic accuracy (AUROC range: 0.698-0.922) depending on the underlying concept, available features, and the applied model technique (Table 3). Model performance increased with the number of available features (*at-the-door* < *collapsed* < *time-series*). Using the same set of features in the *at-the-door* concept, RF and GBM performed similarly with an AUROC of 0.716 and 0.698, respectively. Similar performance differences were found for RF and GBM using additional features in the *collapsed* concept but AUROC and AUPRC results improved compared to the *at-the-door* models (AUROC for RF models: 0.716 vs. 0.873). Best performance was found using LSTM with all available features (*time-series* concept) and time-aware data aggregation (AUROC: 0.922; AUPRC: 0.675).

**Table 3.**
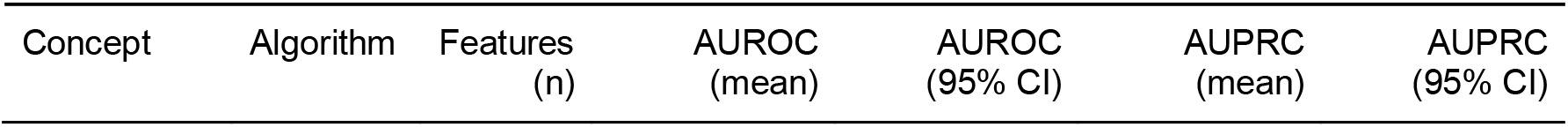
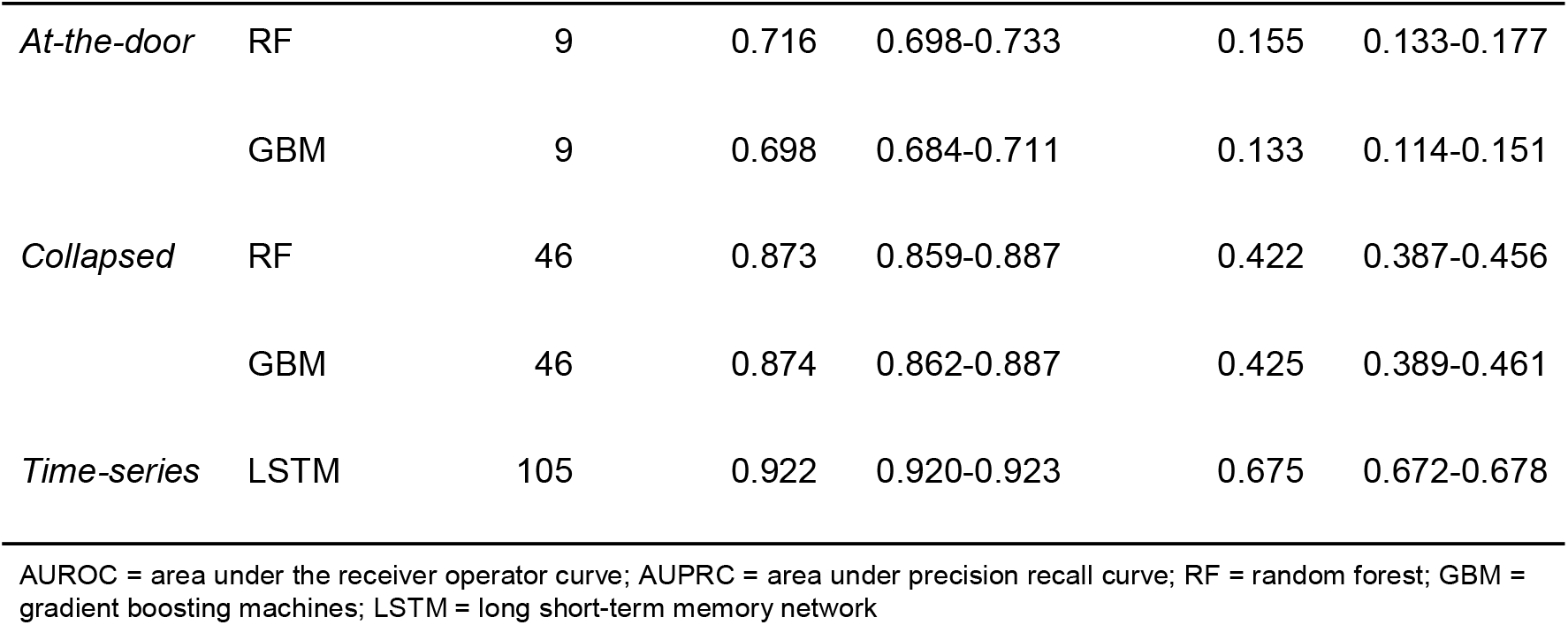
Model performances.

Plotting the AUROC revealed the superiority of the LSTM model (Figure 4). The AUROC also demonstrated the importance of the number of available features while the model technique (RF vs. GBM) resulted in similar performances.

**Figure 4.**
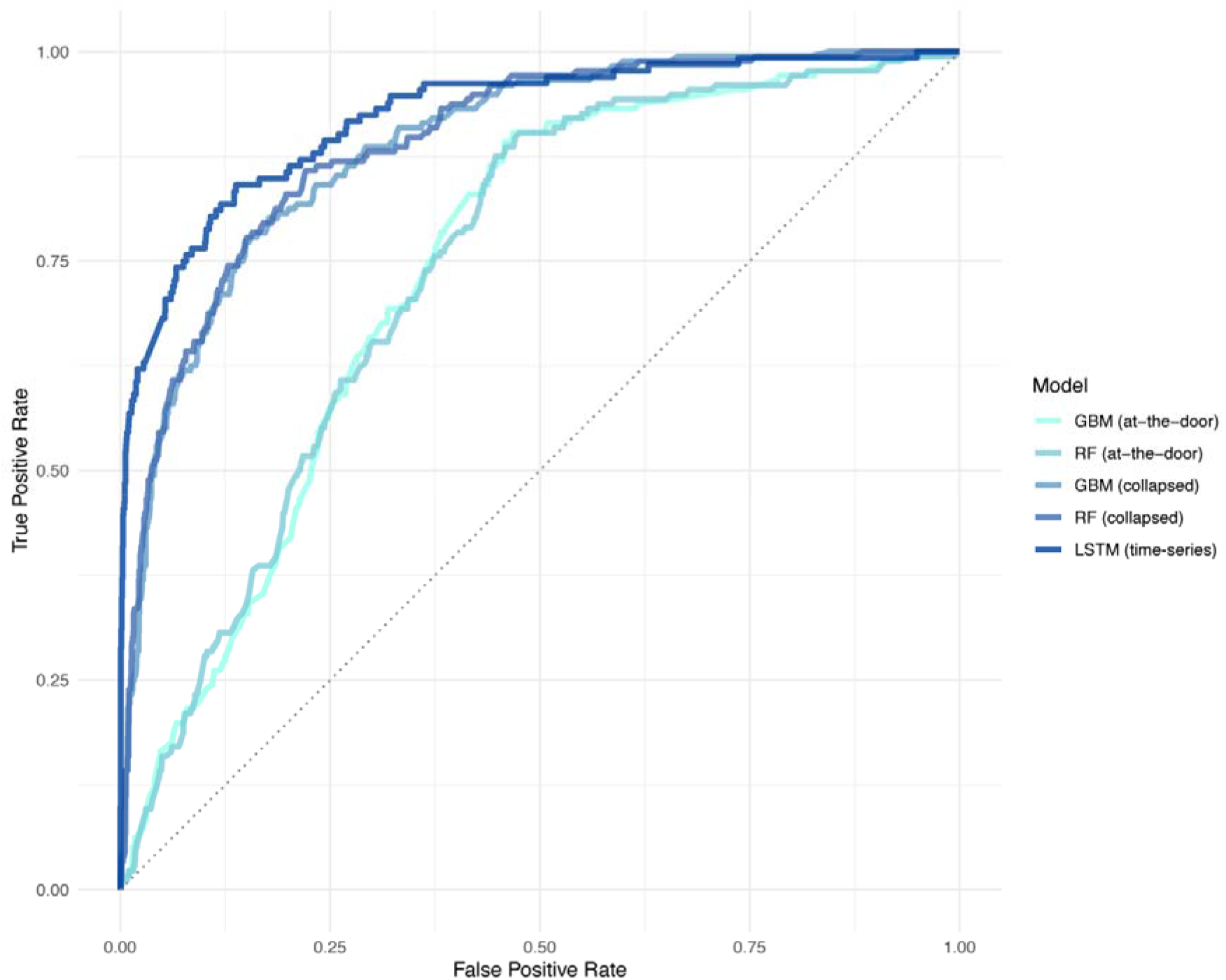
Model performance by area under the receiver operator curve (AUROC) for each model predicting a consultation at the ICU. LSTM showed the highest AUROC of 0.921, reflecting the superiority of the time-series approach.

Given the imbalanced dataset of this study, i.e., the majority of admission (92.2%) did not receive a consultation, assessing the AUPRC provides further information on model performance (Figure 5) [23]. The trade-off between precision (or positive predictive value) and recall (or sensitivity) can be observed and model performance can be compared to the baseline (0.078) which reflects the occurrence of consultations in the study cohort. The LSTM model was also identified to be superior in AUPRC metrics.

**Figure 5.**
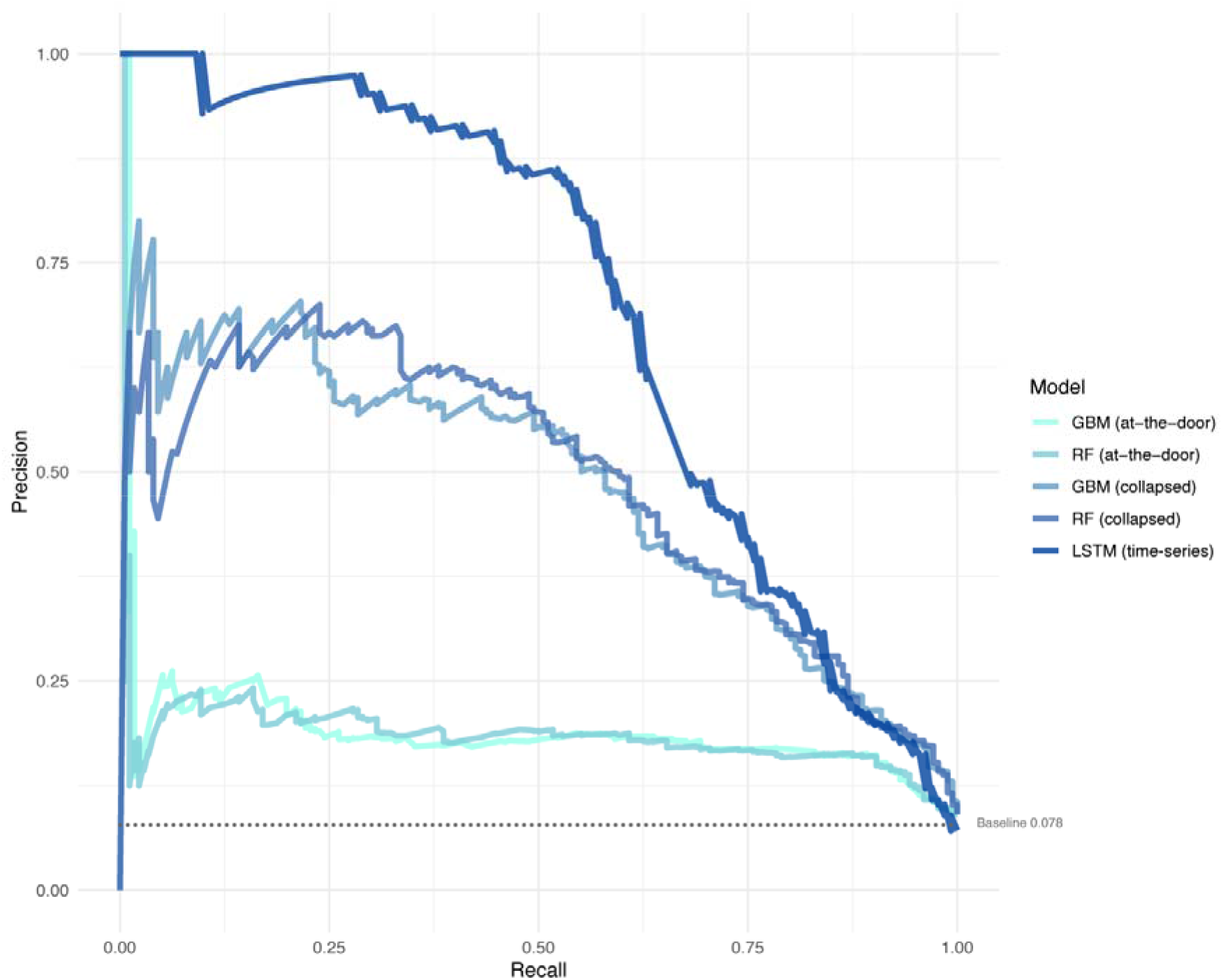
Model performance by area under the precision recall curve (AUCPR) for each model predicting a consultation at the ICU. The baseline represents the performance of a random classifier and reflects the occurrence of consultations in the study cohort (7.8% of all admissions). LSTM showed the highest AUPRC of 0.675, reflecting the superiority of the time-series approach.

## Discussion

This study investigated infection-related consultations (clinical microbiology consultations in this setting) in a cohort of ICU patients and successfully developed a machine learning model for predicting consultations using routine EHR. Early identification of patients requiring a consultation could result in improving optimal allocation and timing of scarce resources, i.e., expert consults, diagnostics, and appropriate antimicrobial treatment. The final model used LSTM, a deep learning technique that can work with time-series data such as EHR. The final dataset comprised 104 features covering demographic, monitoring, procedural, medication, and diagnostic features. The final model performance reached high accuracy with an AUROC of 0.922 and an AUCPR of 0.675.

Infection-related consultations are a relatively rare event for ICU patients and only 7.8% of all admissions in our study cohort received a consultation creating an imbalanced dataset for the target feature. Our cohort included patients that were admitted to the ICU for monitoring purposes, e.g. post-surgery (64.3% in the no-consultation group). These patients typically stay only a short amount of time at the ICU if no complication occurs (median LOS of 2 days in the no-consultation group). Patients with an infection-related consultation also differed significantly from the no-consultation group in age at admission, readmission status, and ICU mortality, thus forming a distinct patient population. The explanatory multiple logistic regression model (Table 2) largely is consistent with empiric clinical reasoning and experience, e.g., the low odds for planned admissions to require a consultation.

Going beyond explanatory modelling by attempting to predict the occurrence of a consultation offers several advantages. Infectious diseases and thus the need for infection-related consultations are highly time-sensitive. Sepsis is one of the most prominent examples often requiring immediate action based on the mere suspicion of sepsis [1]. Initiating diagnostic procedures and adequate treatment, antimicrobial therapy in particular, and the correct and timely ordering of those are essential in the care for critically ill patients. Rapid diagnostics in the (microbiological) laboratory or at point-of-care have evolved greatly over recent years and have the potential to reduce turnaround time substantially, i.e. time from order to clinical action [24,25]. However, technical solutions in the laboratory are costly and do not solve the problem of optimal resource allocation across ICU patients. Potential pre-analytical time gains are often less considered when discussing the concept of rapid diagnostics but the vital aspects of the pre-analytical time and workflow are well established [26]. These considerations also apply to clinical workflows of physicians working in multi-disciplinary teams at the ICU including infection-related consultations. Moreover, our study did not include microbiological data beyond the consultation time stamps and consultations were predicted using only routine ICU data. Thus, one useful case scenario of this prediction model could also be the support of (timely) initiations of microbiological diagnostics.

We demonstrated the successful prediction of consultations in a time shift-based approach with a maximum of eight hours in advance. This could support early identification of patients requiring a consultation and support the initiation subsequent clinical steps (e.g., notifying consultants, performing diagnostics). Our incremental approach showed that an *at-the-door* model with baseline patient characteristics is not sufficient. Although achieving a moderate AUROC, the AUCPR of this model was only little above the baseline in this imbalanced dataset (7.8% of all patients received a consultation). Additional features markedly improved model performance. Using a machine learning technique, LSTM, that can work with time-series data such as EHR was shown to be superior in both AUROC and AUCPR measures. The suitability of LSTM for EHR data was also confirmed in other infection-related studies with similar model performances yet different target events [17–19]. Although model performances are difficult to compare for different outcomes, our results show excellent performance [27]. Moreover, LSTM models offer the advantage of a reduced feature pre-processing necessity.

In clinical practice a time gain of up to eight hours could be beneficial for the patient and all actors involved, intensivists and microbiologists, and help to streamline clinical workflows and allocating resources at the right time for the right patients. The positive impact of infection-related consultations was demonstrated in previous studies as mentioned above [2–6]. First studies on automating consultations also showed promising findings [8,9]. Despite their reactive nature triggered by the occurrence of clinical events, the results pointed towards the feasibility of data-driven consultation workflows. Our proactive approach, i.e., shifting emphasis from reacting to clinical events to predicting an outcome or event, has the potential to bring this to a next level by leveraging existing technology and data.

Our study is subject to limitations. Although we tested our model against a held-out dataset, validation and usability in a real-time scenario need yet to be demonstrated in clinical practice. Machine learning models are known to degrade in performance once deployed and optimal maintenance is important. Our approach might also be affected by this phenomenon. We worked with data that reflect human behaviour (clinical procedures as features, consultation as target outcome). In practice this could influence the model performance as human behaviour might change based on this model’s predictions. In addition, both training and test datasets were used from a single institution and external validation is needed.

However, all features used in the final model are generic features routinely available at ICUs (see Appendix 1). This can facilitate external validation and potential implementation in local EHR systems. Although microbiology reports are also part of routine EHR, they were not included in this study by design. However, the inclusion could yield valuable insights if they influence model performance, which could be explored in future studies. An additional limitation to our approach is the interpretability of the LSTM model. By nature, LSTM and other deep learning techniques do not offer straightforward and interpretable measures such as, for example, feature importance, partial dependence, or individual conditional expectation. If this limitation hinders acceptances by clinicians needs further exploration. We argue that familiarising clinicians with sophisticated machine learning concepts through workflow-centred approaches (e.g., the one presented here) in contrast to more implication-heavy models (e.g., sepsis prediction) could help bridging machine learning technology and development to clinical deployment and acceptance.

## Conclusion

This study demonstrated the feasibility of predicting infection-related consultations for ICU patients up to eight hours in advance. Using routine EHR data and applying a LSTM machine learning model resulted in excellent prediction model performance. In a real-life scenario this approach could help to streamline ICU and clinical microbiology workflows, to allocate scarce resources, and to perform timelier diagnostic and therapeutic interventions in a multi-disciplinary manner. Yet, external validation and further research into acceptance of sophisticated machine learning models by intensivists and consultants is needed. In summary, machine learning supported approaches, such as the one presented here, have great potential to further improve patient care and clinical outcome for critically ill patients with (suspected) infections.

## Data Availability

The data is registered in the Groningen Data Catalogue (https://groningendatacatalogus.nl).

## Acknowledgement

The authors wish to thank Wim Dieperink, Albêrt Heesink, and Martijn Geutjes for their intense support in the data extraction process of this study. We also thank Kim Mouridsen for valuable general discussions.

## Funding

This study was part of a project funded by the European Commission Horizon 2020 Framework Marie Skłodowska-Curie Actions (grant agreement number: 713660-PRONKJEWAIL-H2020-MSCA-COFUND-2015).

## Conflict of Interests

The authors declare no conflict of interests.

## Appendix

**Appendix 1.**
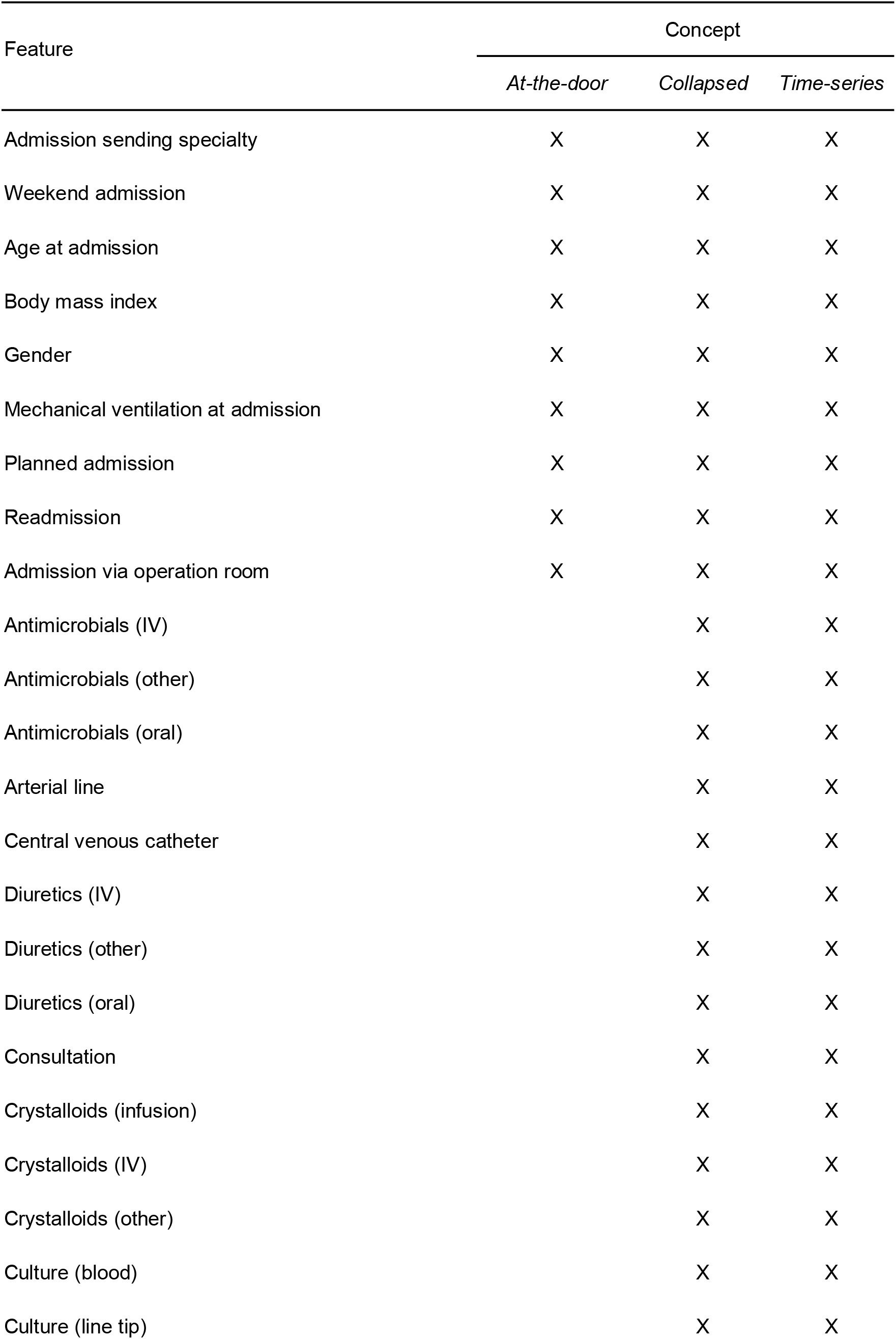
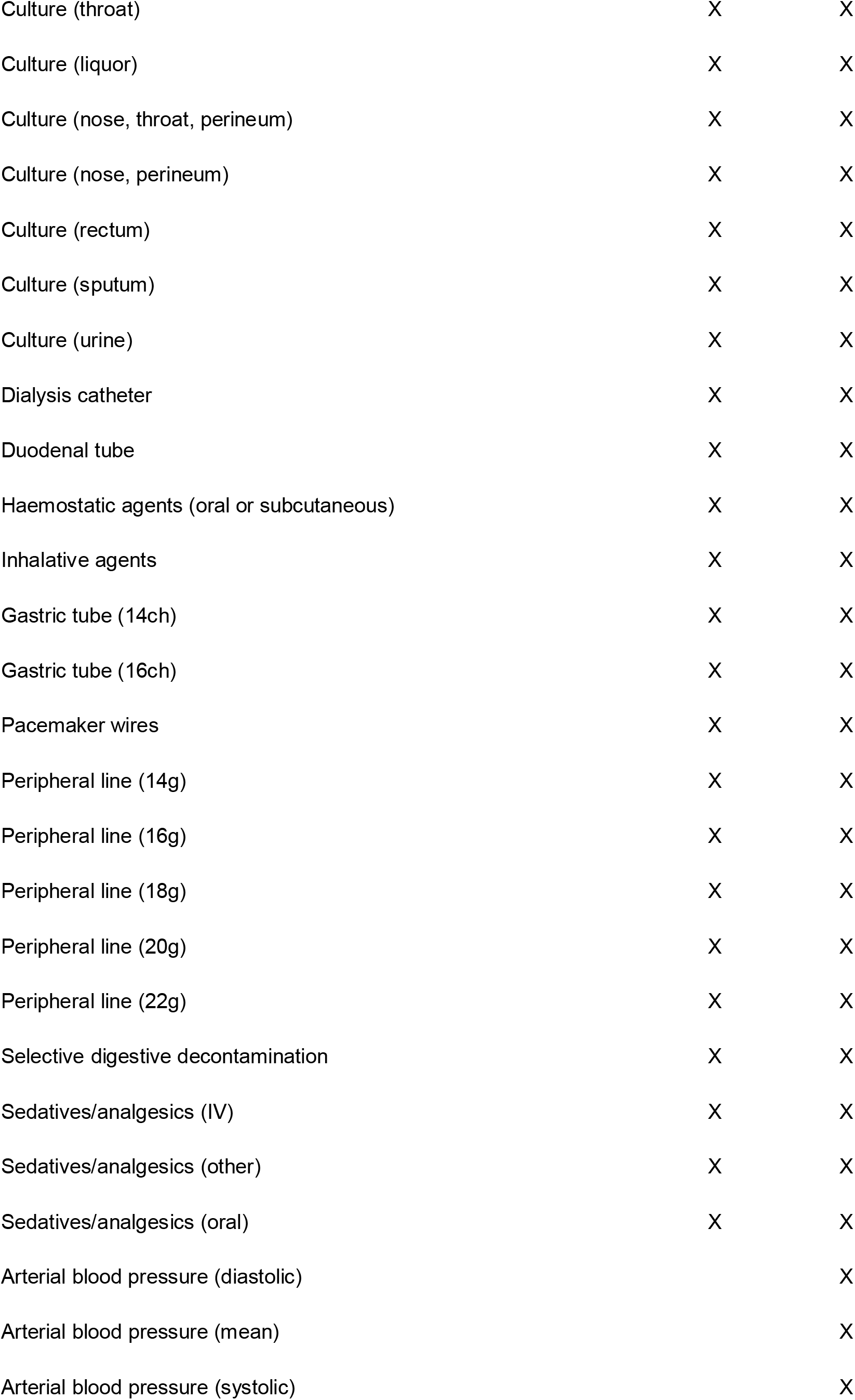
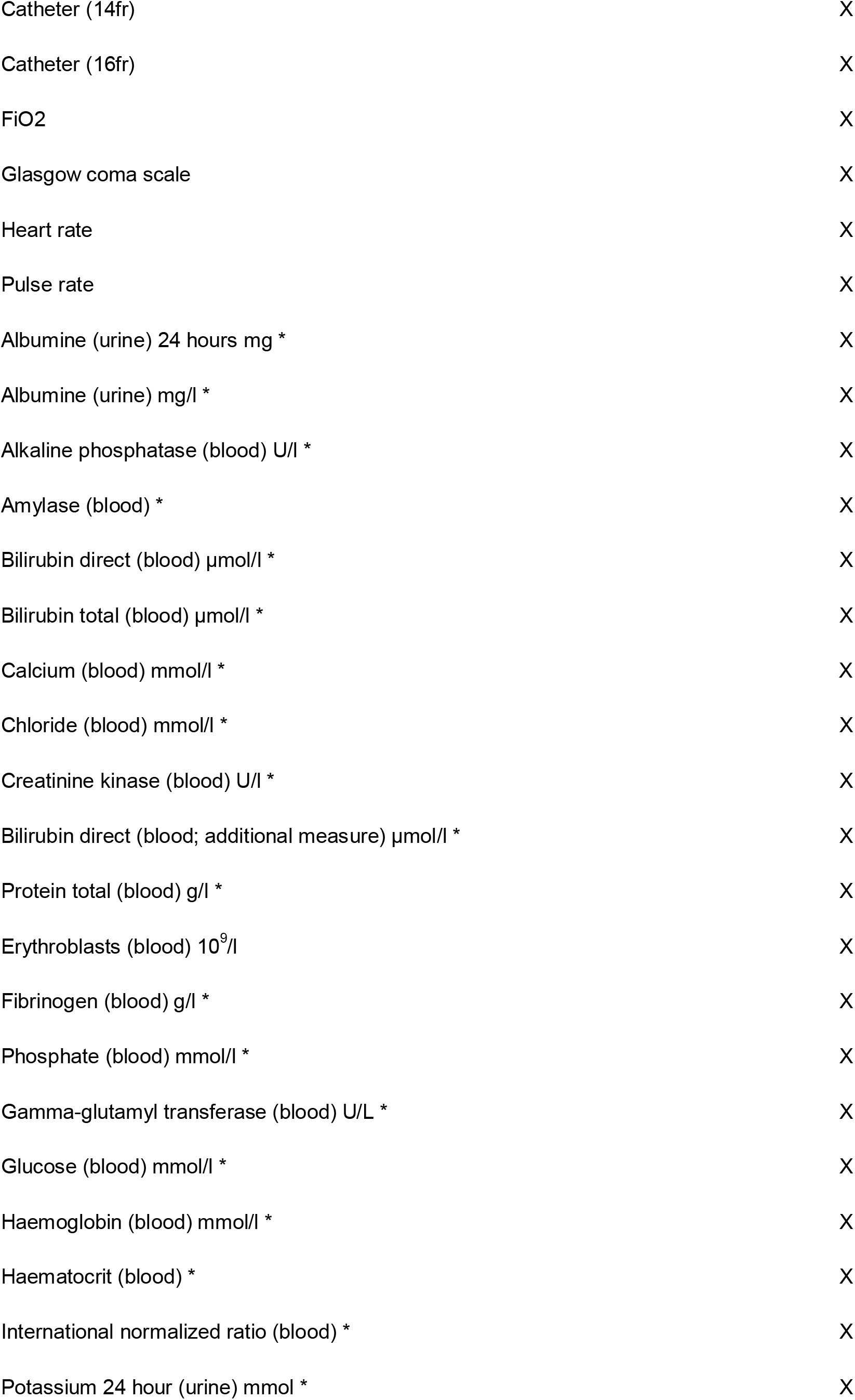
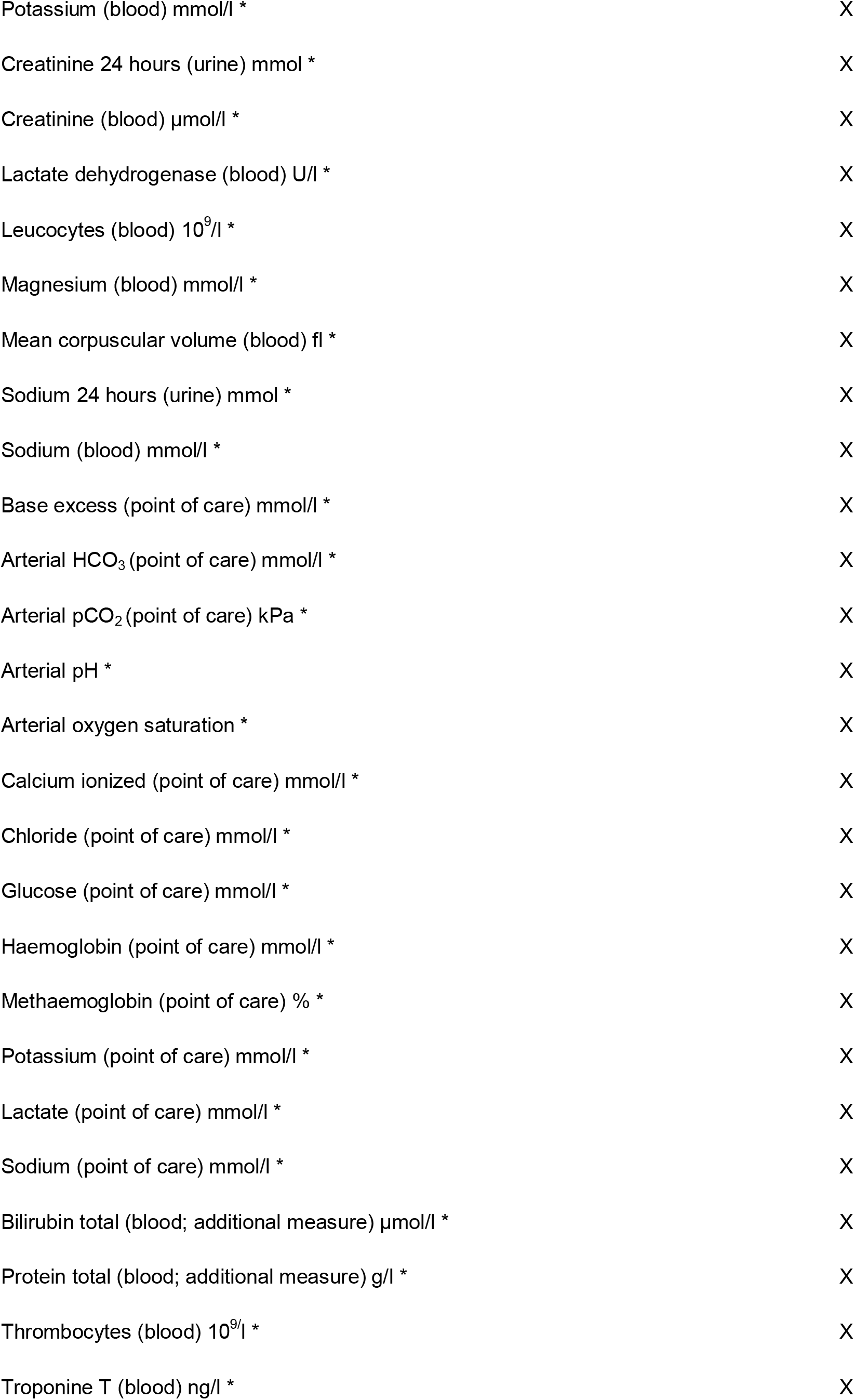
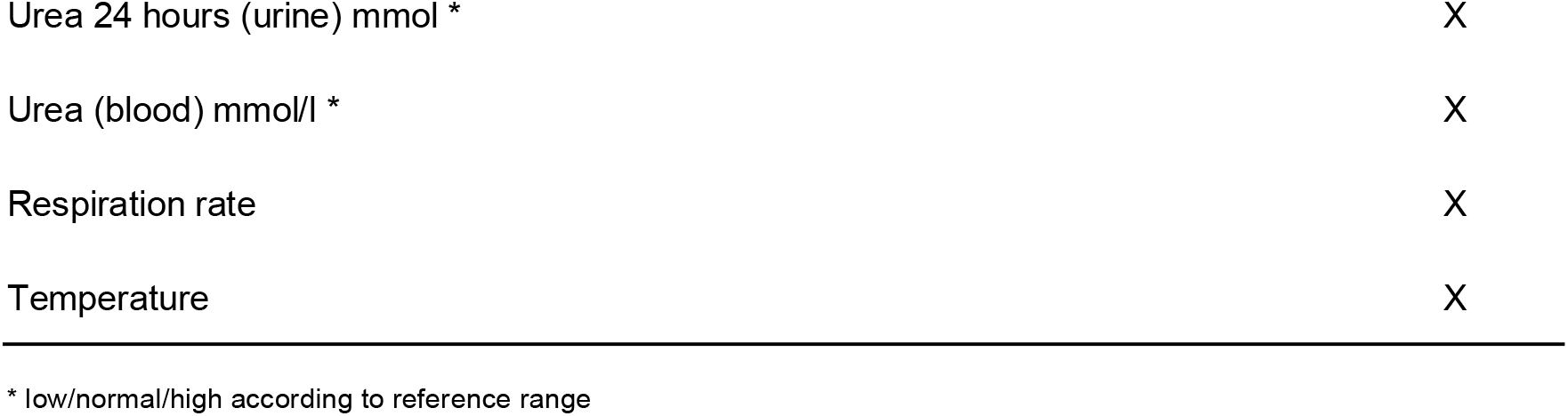
Features used per concept.

## Notes

### Competing Interest Statement

The authors have declared no competing interest.

### Author Declarations

Ethical approval was obtained from the institutional review board of the University Medical Center Groningen and informed consent was waived due to the retrospective observational nature of the study (METc 2018/081).

## References

[1] Rhodes A, Evans LE, Alhazzani W, Levy MM, Antonelli M, Ferrer R, et al. Surviving sepsis campaign: international guidelines for management of sepsis and septic shock: 2016. Crit Care Med 2017;45:486–552. https://doi.org/10.1097/CCM.0000000000002255.

[2] Vogel M, Schmitz RPH, Hagel S, Pletz MW, Gagelmann N, Scherag A, et al. Infectious disease consultation for Staphylococcus aureus bacteremia - A systematic review and meta-analysis. J Infect 2016;72:19–28https://doi.org/10.1016/j.jinf.2015.09.037.

[3] Kampmeier S, Correa-Martinez CL, Peters G, Mellmann A, Kahl BC. Personal microbiological consultations improve the therapeutic management of Staphylococcus aureus bacteremia. J Infect 2018;77:349–56. https://doi.org/10.1016/j.jinf.2018.07.011.

[4] Forsblom E, Ruotsalainen E, Ollgren J, Järvinen A. Telephone consultation cannot replace bedside infectious disease consultation in the management of Staphylococcus aureus Bacteremia. Clin Infect Dis 2013;56:527–35. https://doi.org/10.1093/cid/cis889.

[5] Dik J-WH, Hendrix R, Lo-Ten-Foe JR, Wilting KR, Panday PN, van Gemert-Pijnen LE, et al. Automatic day-2 intervention by a multidisciplinary antimicrobial stewardship-team leads to multiple positive effects. Front Microbiol 2015;6:546. https://doi.org/10.3389/fmicb.2015.00546.

[6] Messacar K, Campbell K, Pearce K, Pyle L, Hurst AL, Child J, et al. A Handshake From Antimicrobial Stewardship Opens Doors for Infectious Disease Consultations. Clin Infect Dis 2017;64:1449–52. https://doi.org/10.1093/cid/cix139.

[7] Stevens JP, Johansson AC, Schonberg MA, Howell MD. Elements of a high-quality inpatient consultation in the intensive care unit. A qualitative study. Ann Am Thorac Soc 2013;10:220–7.https://doi.org/10.1513/AnnalsATS.201212-120OC.

[8] Djelic L, Andany N, Craig J, Daneman N, Simor A, Leis JA. Automatic notification and infectious diseases consultation for patients with Staphylococcus aureus bacteremia. Diagn Microbiol Infect Dis 2018;91:282–3. https://doi.org/10.1016/j.diagmicrobio.2018.03.001.

[9] Jones TM, Drew RH, Wilson DT, Sarubbi C, Anderson DJ. Impact of automatic infectious diseases consultation on the management of fungemia at a large academic medical center. Am J Health Syst Pharm 2017;74:1997–2003. https://doi.org/10.2146/ajhp170113.

[10] Lighthall GK, Vazquez-Guillamet C. Understanding Decision Making in Critical Care. Clin Med Res 2015;13:156–68. https://doi.org/10.3121/cmr.2015.1289.

[11] Gopalan PD, Pershad S. Decision-making in ICU - A systematic review of factors considered important by ICU clinician decision makers with regard to ICU triage decisions. J Crit Care 2019;50:99–110. https://doi.org/10.1016/j.jcrc.2018.11.027.

[12] McKenzie MS, Auriemma CL, Olenik J, Cooney E, Gabler NB, Halpern SD. An Observational Study of Decision Making by Medical Intensivists. Crit Care Med 2015;43:1660–8. https://doi.org/10.1097/CCM.0000000000001084.

[13] DeKeyser Ganz F, Engelberg R, Torres N, Curtis JR. Development of a Model of Interprofessional Shared Clinical Decision Making in the ICU: A Mixed-Methods Study. Crit Care Med 2016;44:680–9. https://doi.org/10.1097/CCM.0000000000001467.

[14] Peiffer-Smadja N, Rawson TM, Ahmad R, Buchard A, Pantelis G, Lescure F-X, et al. Machine learning for clinical decision support in infectious diseases: a narrative review of current applications. Clin Microbiol Infect 2019. https://doi.org/10.1016/j.cmi.2019.09.009.

[15] Luz CF, Vollmer M, Decruyenaere J, Nijsten MW, Glasner C, Sinha B. Machine learning in infection management using routine electronic health records: tools, techniques, and reporting of future technologies. Clin Microbiol Infect 2020. https://doi.org/10.1016/j.cmi.2020.02.003.

[16] de Smet AMGA, Kluytmans JAJW, Blok HEM, Mascini EM, Benus RFJ, Bernards AT, et al. Selective digestive tract decontamination and selective oropharyngeal decontamination and antibiotic resistance in patients in intensive-care units: an openlabel, clustered group-randomised, crossover study. Lancet Infect Dis 2011;11:372–80. https://doi.org/10.1016/S1473-3099(11)70035-4.

[17] Van Steenkiste T, Ruyssinck J, De Baets L, Decruyenaere J, De Turck F, Ongenae F, et al. Accurate prediction of blood culture outcome in the intensive care unit using long short-term memory neural networks. Artif Intell Med 2018. https://doi.org/10.1016/j.artmed.2018.10.008.

[18] Kam HJ, Kim HY. Learning representations for the early detection of sepsis with deep neural networks. Comput Biol Med 2017;89:248–55. https://doi.org/10.1016/j.compbiomed.2017.08.015.

[19] Rajkomar A, Oren E, Chen K, Dai AM, Hajaj N, Hardt M, et al. Scalable and accurate deep learning with electronic health records. Npj Digital Medicine 2018;1:18. https://doi.org/10.1038/s41746-018-0029-1.

[20] R Core Team. R: A Language and Environment for Statistical Computing v3.6.1. Vienna, Austria: 2019.

[21] Python Software Foundation. Python Language Reference, version 3.9. 2020.

[22] Collins GS, Reitsma JB, Altman DG, Moons KGM. Transparent reporting of a multivariable prediction model for individual prognosis or diagnosis (TRIPOD): the TRIPOD statement. BMJ 2015;350:g7594. https://doi.org/10.1136/bmj.g7594.

[23] Saito T, Rehmsmeier M. The precision-recall plot is more informative than the ROC plot when evaluating binary classifiers on imbalanced datasets. PLoS One 2015;10:e0118432. https://doi.org/10.1371/journal.pone.0118432.

[24] Beganovic M, McCreary EK, Mahoney MV, Dionne B, Green DA, Timbrook TT. Interplay between Rapid Diagnostic Tests and Antimicrobial Stewardship Programs among Patients with Bloodstream and Other Severe Infections. J Appl Lab Med 2019;3:601–16. https://doi.org/10.1373/jalm.2018.026450.

[25] Guillamet MCV, Burnham JP, Kollef MH. Novel Approaches to Hasten Detection of Pathogens and Antimicrobial Resistance in the Intensive Care Unit. Semin Respir Crit Care Med 2019;40:454–64. https://doi.org/10.1055/s-0039-1693160.

[26] Lamy B, Sundqvist M, Idelevich EA, ESCMID Study Group for Bloodstream Infections, Endocarditis and Sepsis (ESGBIES). Bloodstream infections - Standard and progress in pathogen diagnostics. Clin Microbiol Infect 2020;26:142–50. https://doi.org/10.1016/j.cmi.2019.11.017.

[27] Hosmer DW Jr, Lemeshow S, Sturdivant RX. Applied Logistic Regression. Hoboken: John Wiley & Sons; 2013.

